# Intrahost-diversity of influenza A virus in upper and lower respiratory tract derived samples from a college community

**DOI:** 10.1101/2021.10.27.21265424

**Authors:** Nicolae Sapoval, P. Jacob Bueno de Mesquita, Yunxi Liu, Roger Wang, Tian Rui Liu, Josie Garza, Torrey Williams, Carmelli Cadiz, Gene S. Tan, Harm Van Bakel, R. A. Leo Elworth, Michael L. Grantham, EMIT Investigators, Donald K. Milton, Todd J. Treangen

**Affiliations:** Department of Computer Science, Rice University, Houston, TX 77005; Public Health Aerobiology and Biomarker Laboratory, Institute for Applied Environmental Health, University of Maryland School of Public Health, College Park, MD 20742; Infectious Diseases, J. Craig Venter Institute, La Jolla, CA 92037; Division of Infectious Diseases, Department of Medicine, University of California San Diego, La Jolla, CA 92037; Department of Genetics and Genomic Sciences, Icahn School of Medicine at Mount Sinai, New York, NY 10029, USA; Icahn Institute for Data Science and Genomic Technology, Icahn School of Medicine at Mount Sinai, New York, NY 10029, USA; Department of Biology, Missouri Western State University, Saint Joseph, MO 64507, USA

## Abstract

**Motivation:** Influenza is a rapidly mutating RNA virus responsible for annual epidemics causing substantial morbidity, mortality, and economic loss. Characterizing influenza virus mutational diversity and evolutionary processes within and between human hosts can provide tools to help track and understand transmission events. In this study we investigated possible differences between the intrahost genomic content of influenza virus in upper respiratory swabs and exhaled aerosols thought to be enriched for virus from the lower respiratory tract.

**Results:** We examined the sequences of specimens collected from influenza A virus (IAV) infected college community members from December 2012 through May 2013. We analyzed four types of IAV samples (fine ≤5 µm aerosols (N=38), coarse >5µm aerosols (N=27), nasopharyngeal (N=53), and oropharyngeal swabs (N=47)) collected from 42 study participants with 60 sampling instances. Eighteen (42.9%) participants had data from four sample types (nasopharyngeal swab, oropharyngeal swab, coarse aerosol, fine aerosol) included in the analysis, 10 (23.8%) had data from 3 sample types, 10 (23.8%) had data from 2 sample types, and 4 (9.5%) had data from one type of sample included in the analysis. We found that 481 (53.3%) consensus single nucleotide polymorphisms are shared by all sample types and 600 (66.5%) are shared by at least three different sample types. We observed that within a single patient consensus and non-consensus single nucleotide variants are shared across all sample types. Finally, we inferred a phylogenetic tree using consensus sequences and found that samples derived from a single patient are monophyletic.

**Conclusions:** Single nucleotide polymorphisms did not differentiate between samples with varying origin along the respiratory tree. We found that signatures of variation in non-consensus intrahost single nucleotide variants are host and sample, but not site specific. We conclude that the genomic information available does not allow us to discern a transmission route. Future investigation into whether any site-specific mutational signatures emerge over a longer period of infection, for example in immunocompromised hosts, can be interesting from the virus evolution perspective.

## INTRODUCTION

The risk of influenza transmission by aerosols versus contact and spray-borne transmission remains uncertain despite its importance to the scientific underpinnings of population disease control and prevention (Cowling et al. 2013; Killingley and Nguyen-Van-Tam 2013; Weinstein et al. 2003). A human-challenge transmission trial – thought to be the most promising approach to address this question – achieved minimal transmission and suggested an important role of aerosols without the ability to confidently quantify aerosol risk (Bueno de Mesquita et al. 2020; Nguyen-Van-Tam et al. 2020). While possible to improve on the human-challenge transmission trial model, the observational study of wildtype influenza infection and evolution in the community, as has been done in household studies, represents a promising approach with perhaps greater generalizability to a variety of human populations and viral communities. Tracing acute respiratory infections by transmission mode could help identify epidemiologic features of transmission events including possible high risk transmission environments (e.g., poorly ventilated indoor spaces) (Delikhoon et al. 2021) and characteristics of super-spreaders (Stein 2011), thus, illuminating the potential benefit of airborne control measures (Morawska et al. 2020). Minor-variants have been used to help infer influenza and other viruses transmission chains (Stack et al. 2013; Park et al. 2015; Poon et al. 2016; McCrone et al. 2018; Müller et al. 2020; Berry et al. 2021) and evolutionary dynamics (Pybus and Rambaut 2009; Rodriguez-Roche et al. 2016; Kijak et al. 2017). Further variation in mutational profiles between sample types within individuals could provide additional information to refine the reconstruction of transmission chains. This refinement has the potential to point out the infection route between individuals with sequenced aerosol and upper respiratory mucosal specimens. However, a precursor to the use of genomic variant data to confidently identify transmission chains by mode is whether there exists enough distinguishable genomic variation in viruses shed from various sites along the respiratory tree during infection.

Bottleneck size has been postulated as a feature that may differentiate airborne versus contact transmission modes (Varble et al. 2014; Frise et al. 2016; Sigal et al. 2018). A smaller influenza viral bottleneck was observed in susceptible ferrets receiving an aerosol versus close contact exposure to infected ferrets where aerosol and/or direct contact modes could have occurred (Frise et al. 2016). A similar ferret study reported a small aerosol bottleneck size but found that viral communities in the lower respiratory tract may not be as likely to transmit infection via aerosols as upper respiratory communities (Varble et al. 2014). This may represent an important difference in comparison with humans where viral loads in the nose and the lung of symptomatic influenza cases were found to be uncorrelated (Yan et al. 2018) and viral shedding into aerosols from the distal lung may pose an important source of transmission for humans (Tellier et al. 2019).

Genetically confirmed household transmission pairs, with unresolved transmission modes, were identified, with an estimated transmission bottleneck size of 1-2 genomes for influenza A virus (McCrone et al. 2018). Bioinformatics methods have been developed to synthesize pathogen respiratory viral genomic sequence data with epidemiological contact and exposure information to confirm whom transmitted to whom (Didelot et al. 2014; Jombart et al. 2014b, 2014a; Campbell et al. 2019). Additionally, minor-variant analysis has been used to infer influenza transmission networks (Stack et al. 2013; Poon et al. 2016; McCrone et al. 2018). But the extent to which transmission pairs can be further differentiated by mode of infection is unclear. Influenza A H1N1 viral variants associated with moderate to severe influenza-like illness were more likely to be detected in the lower respiratory tract from bronchoalveolar lavage compared with upper respiratory nasal swabs (Piralla et al. 2011). Furthermore, it has been shown that influenza A virus has overall higher genomic diversity than influenza B virus (Valesano et al. 2020). It remains uncertain whether viral variants of lower respiratory tract origin collected in exhaled breath of influenza cases differ from viral variants from regions higher in the respiratory tree detected in oropharyngeal and nasopharyngeal swabs.

We sequenced specimens from symptomatic, naturally infected influenza cases from a college community to investigate whether independent sets of influenza variants might exist between upper and lower respiratory specimens. We characterized non-synonymous variants and variant stability over the course of up to three days post symptom onset, as well as the frequency of consensus single nucleotide polymorphisms (SNPs) and intrahost single nucleotide variants (iSNVs) differentiating samples from the upper and lower respiratory tracts. Additionally, we have also investigated the potential for identifying upper or lower airways as the source of viral transmission.

## METHODS

### Study population and sample collection

Samples were taken from the EMIT study of symptomatic, naturally infected influenza cases at the University of Maryland (2012-2013). The study design and methods have been previously reported (Yan et al. 2018). Briefly, the study was reviewed and approved by the University of Maryland Institutional Review Board. Individuals presenting to the campus health center or to the research clinic with influenza-like illness were invited to participate in the study. Volunteers were screened and enrolled if they were within the first 3 days of symptom onset and met the study’s influenza case definition: (1) positive Quidel QuickVue influenza A or B test, or (2) oral temperature >37.8°C and reported having a cough or sore throat. Volunteers answered a questionnaire about symptoms, vaccination, and medical history but a systematic investigation of contacts was not conducted. Samples were collected on up to three consecutive days post symptom onset. Fine (≤5 µm) and coarse (>5µm) exhaled breath aerosols oropharyngeal and nasopharyngeal swabs (NPS) were collected and processed as previously described. Aerosol and NPS samples were evaluated via quantitative reverse-transcriptase polymerase chain reaction (qRT-PCR) and viral culture. All samples were stored at −80°C until sequencing. Respiratory and systemic symptom scores, smoker and asthma status, and vaccination history were also documented.

### Extraction and sequencing methods

All samples were extracted and amplified using the sequence independent single primer amplification (SISPA) method and sequenced using a MiSeq 2×300 bp at the J. Craig Venter Institute for genomic research. To increase depth of coverage, remaining RNA and original sample material were sent to Mt. Sinai for resequencing (with extraction step if using original sample material). Sample extractions were performed using the standard Qiagen viral RNA spin protocol. Following RNA extraction, a multi-segment one-step reverse-transcriptase PCR was performed to generate virus-specific amplicons (Zhou et al. 2009). A second amplification of the viral amplicons was performed using the SISPA (Djikeng et al. 2008); this was done twice for each sample (double barcoded). Samples were then sequenced using an Illumina MiSeq machine (2 × 300 bp).

### Sequencing samples and data available for analysis

The final set of samples with sufficient sequence data for analysis included 60 sampling instances from 42 study participants with qRT-PCR confirmed influenza viral infection. We included in the analysis all samples with sufficient sequence data, including samples from 18 (42.9%) participants with four sample types (nasopharyngeal swab, oropharyngeal swab, coarse aerosol, fine aerosol), 10 (23.8%) participants with 3 sample types, 10 (23.8%) with 2 sample types, and 4 (9.5%) with one sample type. The population was generally young adult and healthy, with illnesses characterized by substantial observed coughing and systemic illness (Table 1). Fifteen of the 42 participants (36%) had data on more than one day during acute illness.

**Table 1.** Sociodemographic, health history, and acute symptoms of study population

| Variable | Median (IQR) or Frequency (%)<br>(N=42 participant-infections) |
| --- | --- |
| Age | 20.5 (19.0, 21.0) |
| Male sex | 20 (47.6%) |
| BMI | 23.5 (21.2, 26.3) |
| Asthma | 8 (19.0%) |
| Smoker | 3 (7.1%) |
| Vaccine for current season | 10 (23.8%) |
| Vaccination for current and previous season | 8 (19.0%) |
| Asymptomatic | 0 (0.0%) |
| Fever > 37.9°C | 6 (14.3%) |
| Variable | N = 60 sampling instances<br>(up to 3 days of of sampling per participant-infection) |
| Antiviral use within last 24 hr | 10 (5.8%) |
| Oral temperature | 37.3 (37.0, 37.6) |
| Cough count | 29.0 (14.0, 52.0) |
| Sneeze count | 0.0 (0.0, 0.0) |
| Upper respiratory score | 7.0 (5.0, 9.0) |
| Lower respiratory score | 3.5 (3.0, 6.0) |
| Systemic symptom score | 9.0 (6.0, 10.0) |
| Total symptom score | 20.0 (15.0, 23.0) |

IQR: Interquartile range. Fever >37.9°C during any day of observation through acute illness. Number of coughs or sneezes counted during 30 minute exhaled breath collection. Asymptomatic: no symptoms at all during acute illness (unlikely due to study inclusion based on symptoms and rapid-test positive or fever plus cough or sore throat). Upper respiratory score: sum of runny nose, stuffy nose, sneeze, sore throat, earache; Lower respiratory score: sum of chest tightness, shortness of breath, cough; Systemic symptom score: sum of malaise, headache, muscle and joint ache, sweats/fever/chills, where each individual symptom score ranges ordinally from 0 (no symptom at all) to 3 (severe symptom). Three (7.1%) participants had data from at least one sample type on 3 days of follow up, 12 (28.6%) had data from 2 days of sampling, and 27 (64.3%) had data from 1 day of sampling.

### Read mapping and variant calling methods

The sequencing reads were pre-processed for adapter removal and quality control purposes using trimmomatic v0.39 with a set of universal adapters (trimmomatic PE ILLUMINACLIP:{adapter_file}:2:30:10 LEADING:3 TRAILING:3 SLIDINGWINDOW:4:15 MINLEN:36) (Bolger et al. 2014). The trimmed reads were mapped to Flu genome segments (NCBI reference sequence NC_007366.1-NC_007373.1) with bowtie2 v2.3.5 (bowtie2 --local --maxins 600) (Langmead and Salzberg 2012). SAM files were converted to BAM and sorted with samtools v1.11 (samtools view -bS, samtools sort -O BAM, samtools index) (Li et al. 2009). Variant calling was done using LoFreq v2.1.4 with default parameters (lofreq call-parallel) (Wilm et al. 2012).

## RESULTS

We present results focusing on the identification of within-person viral variants in NP swabs, oropharyngeal swabs and coarse and fine particle aerosols using influenza-specific deep sequencing.

### Mutational spectra of samples collected from upper and lower respiratory tracts are similar

Our results show that the SNP mutational profile (Figure 1) is consistent for all sample types. In the case of iSNVs we observe greater variability in the mutational profiles (Figure 2), indicating that iSNVs can help differentiate different sample types of the influenza virus. Furthermore, we note that most of the SNP calls in the influenza data are shared between different sample types (481 SNPs; Figure 3) while a small number of SNPs occurred only in a single sample type (coarse aerosols: 28 SNPs, fine aerosols: 28 SNPs, nasopharyngeal: 26 SNPs, oropharyngeal: 7 SNPs; Figure 3). iSNV calls on the other hand appear to be much more sample type specific, with only a single iSNV being shared across all sample types (Figure 4).

**Figure 1.**
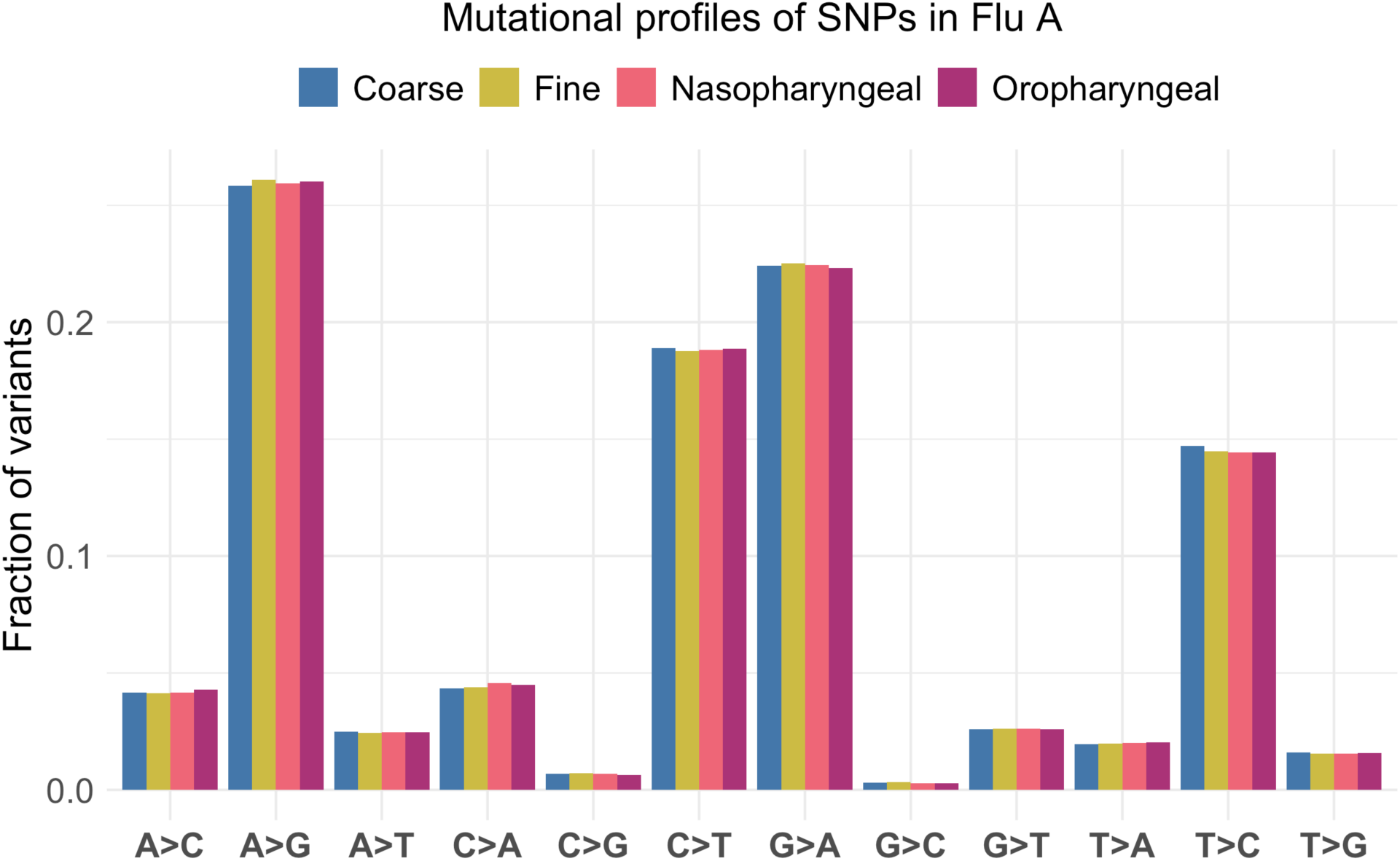
Mutational profile of SNPs (AF > 50%) in influenza A data. Different sample types are represented with different colors and organized left to right per SNP type (coarse aerosols = blue, fine aerosols=yellow, nasopharyngeal swabs = pink, oropharyngeal swabs = purple).

**Figure 2.**
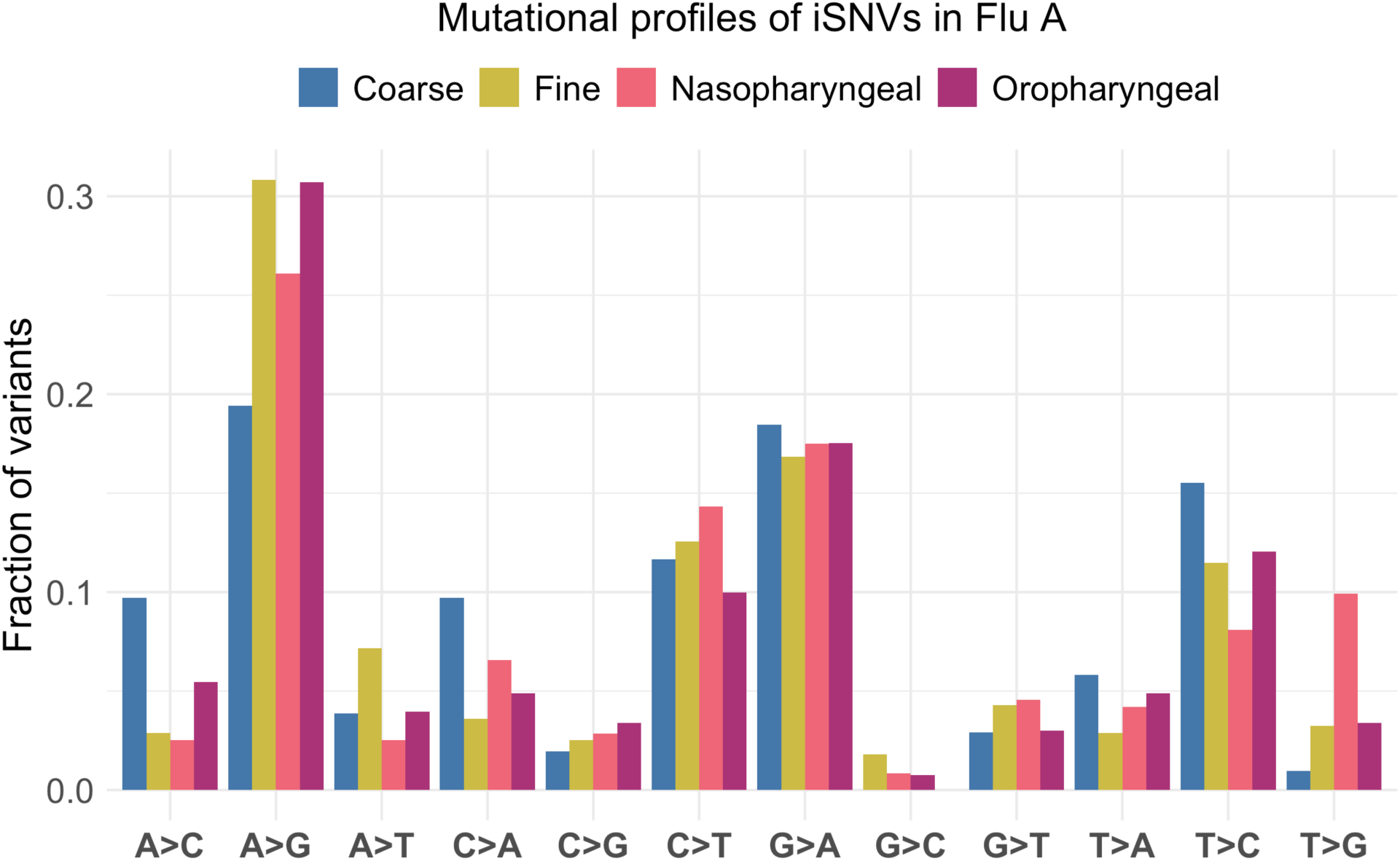
Mutational profile of iSNVs (AF < 50%) in influenza A data. Different sample types are represented with different colors and organized left to right per iSNV type (coarse aerosols = blue, fine aerosols=yellow, nasopharyngeal swabs = pink, oropharyngeal swabs = purple).

**Figure 3.**
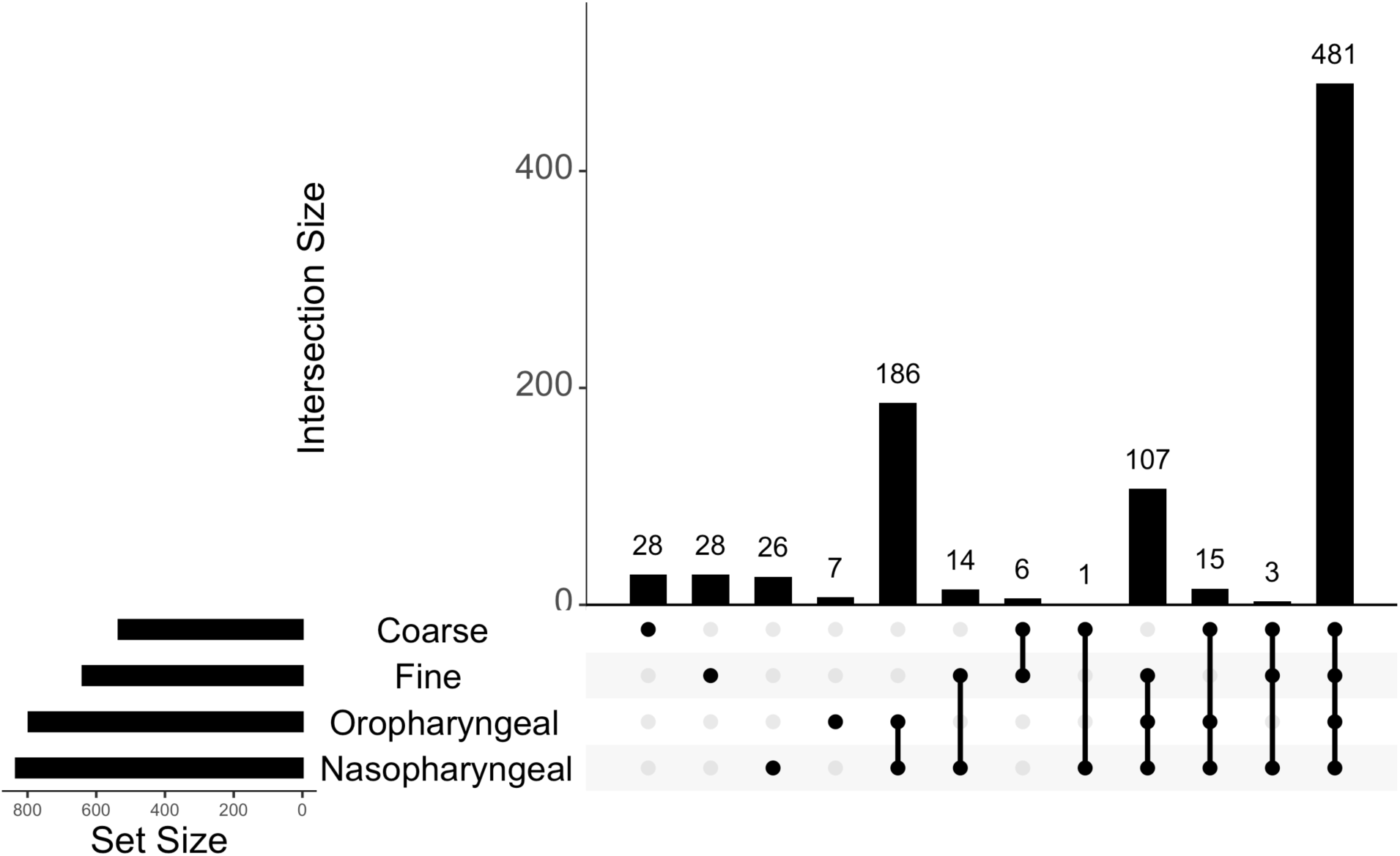
UpSet plot capturing overlaps between SNPs within the influenza A dataset. Bars represent the size of the overlap, and the dots represent the SNP sets that are being overlapped.

**Figure 4.**
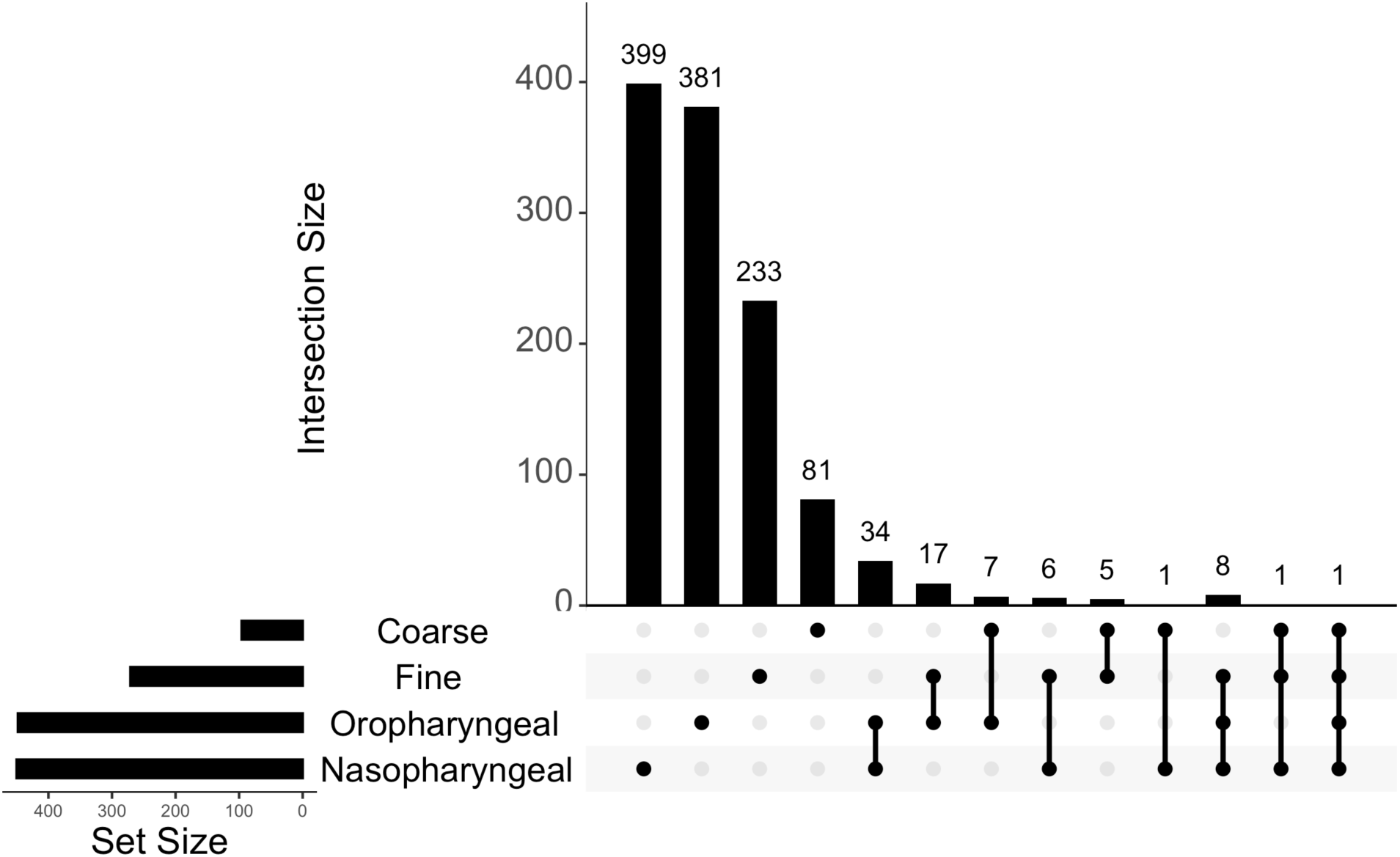
UpSet plot capturing overlaps between iSNVs within the influenza A dataset. Bars represent the size of the overlap, and the dots represent the iSNV sets that are being overlapped.

### Stability of mutational profiles for up to three days post onset of symptoms

For study participants that had data over multiple days, as well as data from different sample types (two participants in total, subject 63 and subject 79), we compared the variant stability over time by pooling the iSNVs and SNPs and examining overlap within sample types (Figures 5&6). For subject 63, we had 11 samples (3 nasopharyngeal, 3 oropharyngeal, 3 fine aerosol, and 2 coarse aerosol) collected over the course of 3 days. For subject 79, we had 10 samples (2 nasopharyngeal, 3 oropharyngeal, 3 fine aerosol, and 2 coarse aerosol) collected over the course of 3 days. We note that nasopharyngeal and oropharyngeal samples exhibit longitudinal stability when viewing the variants over the course of infection (Figure 5A, B and Figure 6A, B). Fine and coarse aerosol samples show less consistent behavior. In subject 79 we observe variant stability over the course of the infection within fine aerosol but not the coarse aerosol samples (Figure 6C, D), while in subject 63 there is a notable variability in the variants detected in both aerosol fractions (Figure 5C, D). Additionally, when we compare the pooled iSNV and SNP stability between different sample types within the same subject (Figure 7) we note that for both subjects most of the variants are common to all sample types. These observations indicate that most variants within the single participant converge to the same set over the course of infection.

**Figure 5.**
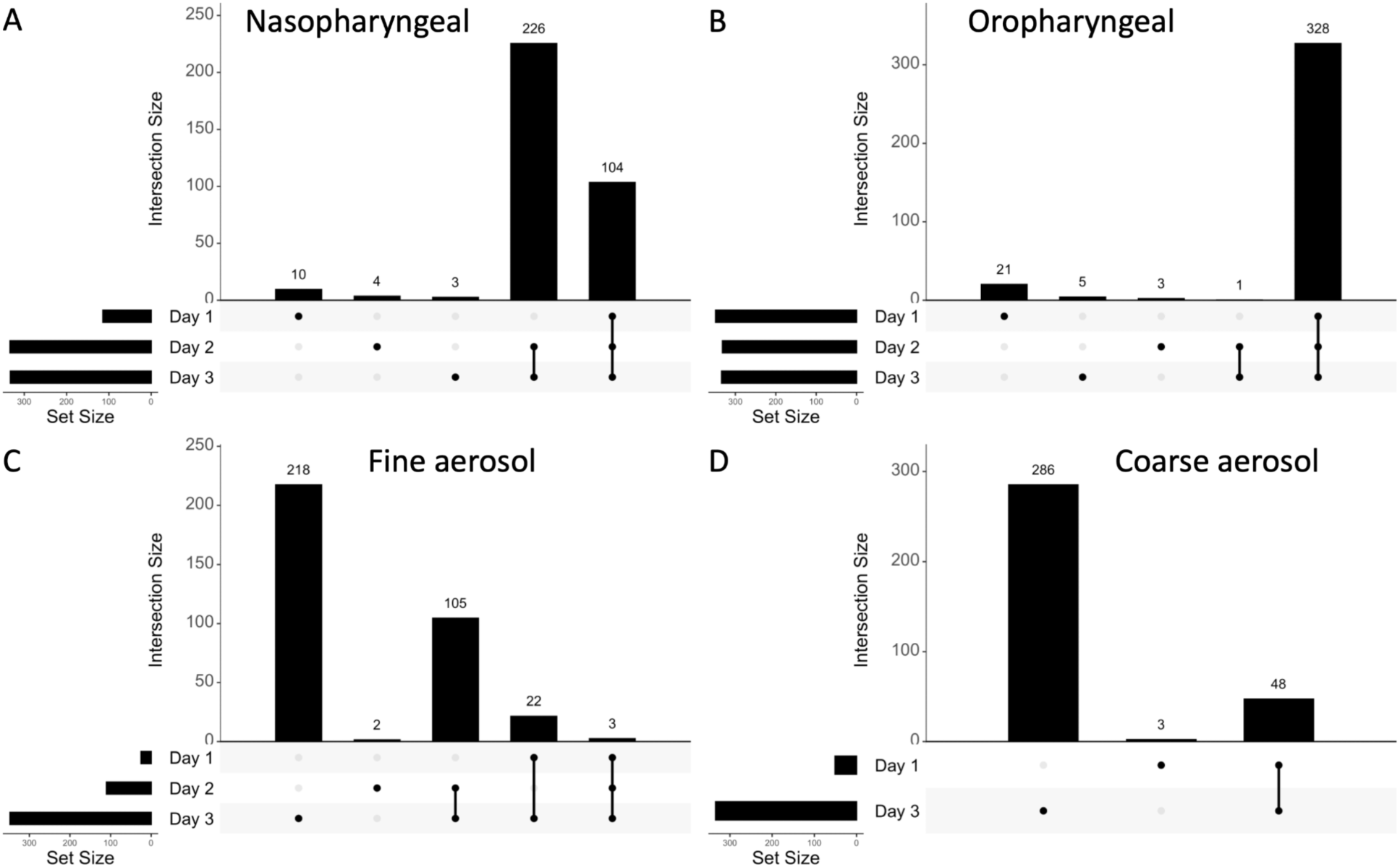
UpSet plots capturing overlaps among pooled iSNVs and SNPs within sample type over time for study subject 63. Bars represent the size of the overlap, and the dots represent the iSNV sets that are being overlapped. (A) Nasopharyngeal data over the course of 3 days past onset. (B) Oropharyngeal data over the course of 3 days past onset. (C) Fine aerosol data over the course of 3 days past onset. (D) Coarse aerosol data on the 1st and 3rd days past onset.

**Figure 6.**
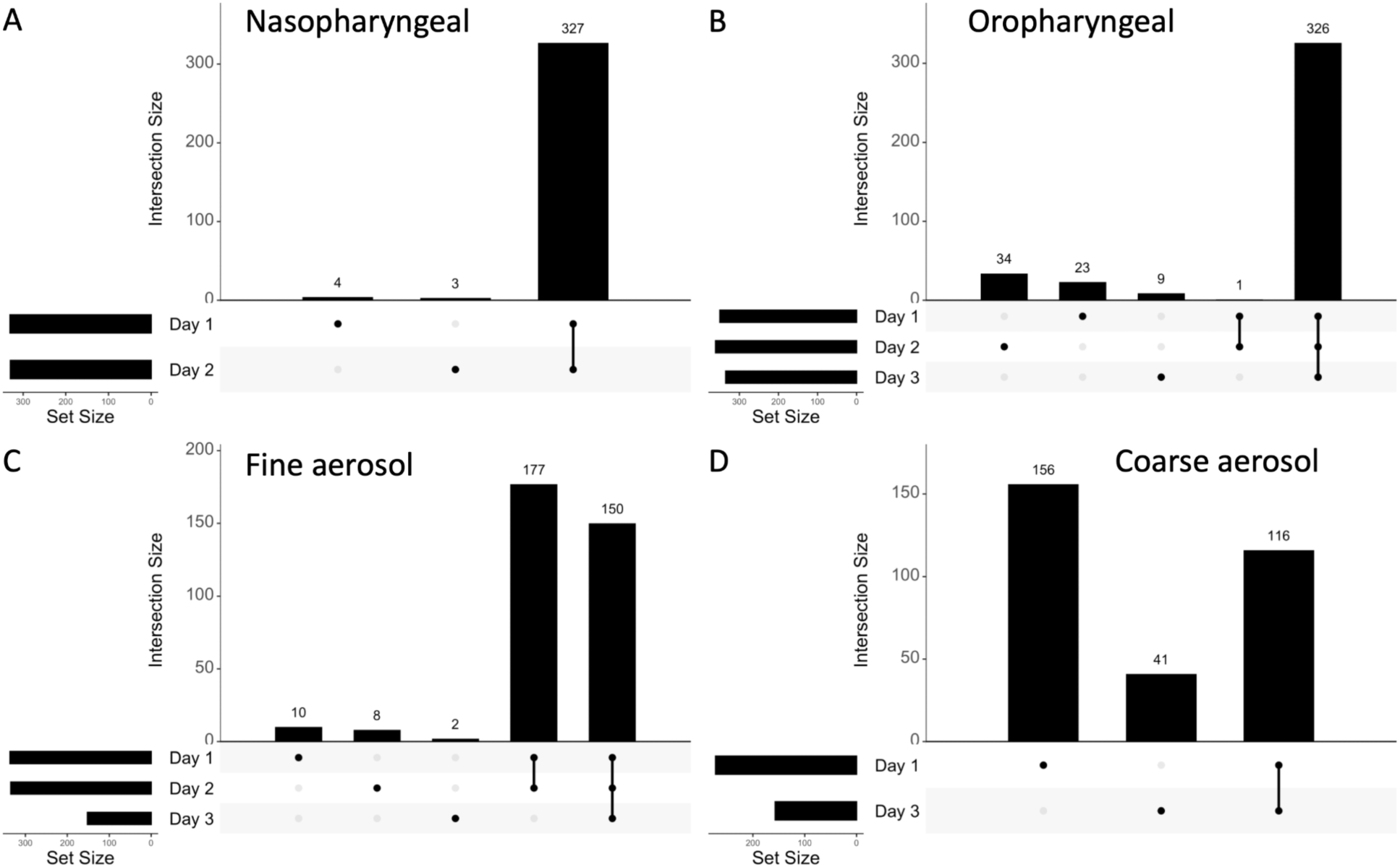
UpSet plots capturing overlaps among pooled iSNVs and SNPs within sample type over time for study subject 79. Bars represent the size of the overlap, and the dots represent the iSNV sets that are being overlapped. (A) Nasopharyngeal data over the course of 2 days past onset. (B) Oropharyngeal data over the course of 3 days past onset. (C) Fine aerosol data over the course of 3 days past onset. (D) Coarse aerosol data on the 1st and 3rd days past onset.

**Figure 7.**
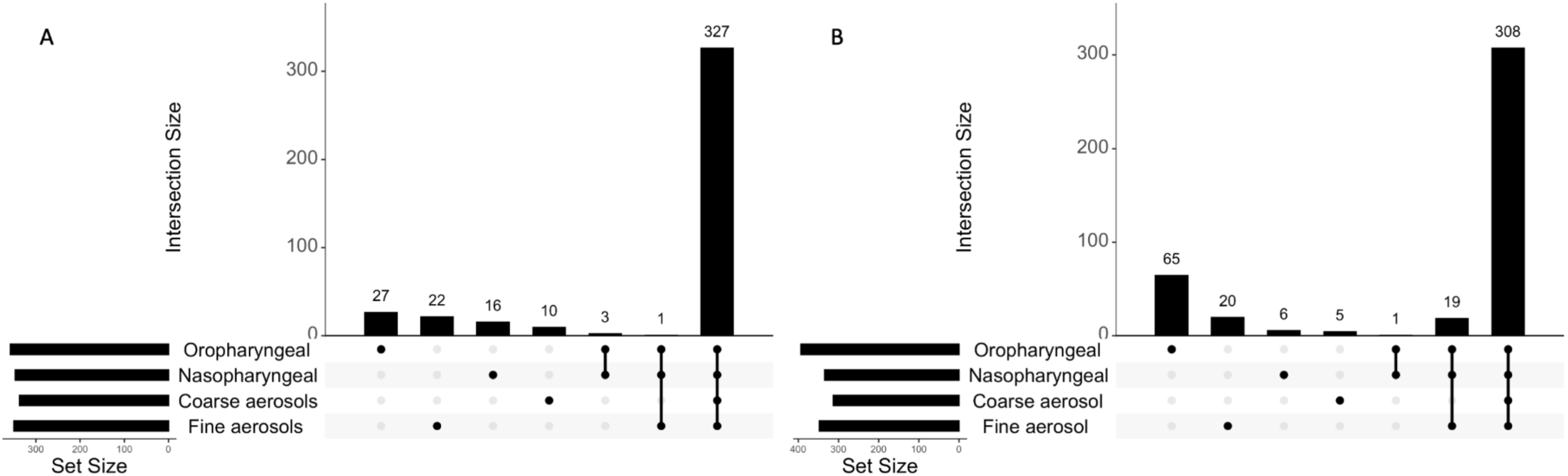
UpSet showing the overlaps among pooled iSNVs and SNPs based on the sample type for study subject 63 (A) and 79 (B) over the 3 days post symptom onset.

We have annotated the SNPs identified in this study based on their effect and compared the profiles of missense and synonymous mutations across SNPs that were only found in a specific sample type (Figure 8). We observe that in all cases except for the oropharyngeal swab samples (which have less than 10 sample type specific SNPs in total) more than half of all SNPs are synonymous mutations (Figure 8A, C, D).

**Figure 8.**
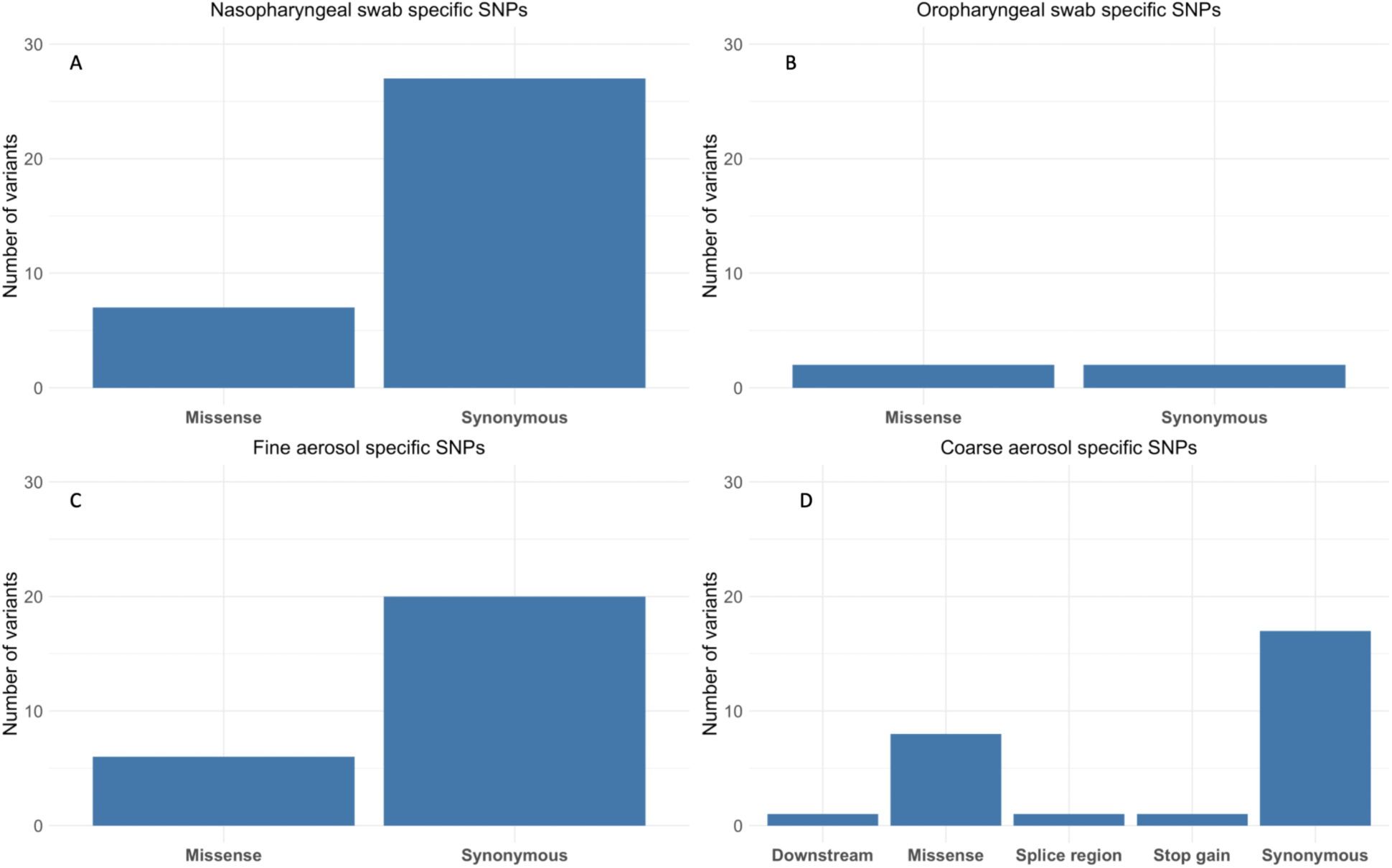
Characterization of SNPs specific to each of the sample types based on the mutation effect. (A) SNPs specific to nasopharyngeal samples. (B) SNPs specific to oropharyngeal samples. (C) SNPs specific to fine aerosol samples. (D) SNPs specific to coarse aerosol samples.

### Phylogenetic inference on the influenza samples

We have inferred a phylogenetic tree consisting of joined segments of the Flu A virus (Figure 9) with samples being represented by the respective consensus sequences. We note that most samples derived from the same participant are monophyletic, with most of consensus sequences for these samples being identical (indicated by the same color). We also note that most branches are short, except for the clade containing participants 48 and 113. This feature of the tree points to the possibility of a single introduction event being the primary source of spread for the flu virus within this community. Due to the lack of the strong phylogenetic signal at the consensus level (SNPs) and longitudinal instability of the majority of iSNVs, we were unable to fully investigate the potential for inferring transmission route between subjects.

**Figure 9.**
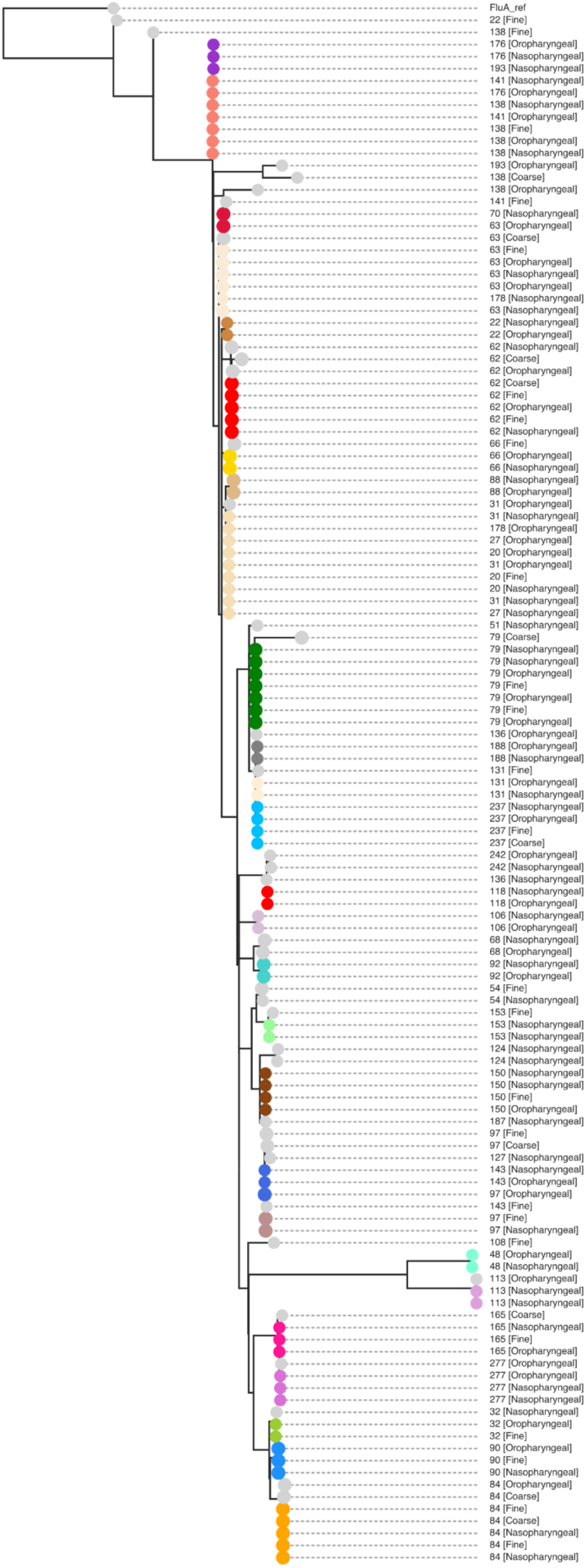
Inferred phylogenetic tree for the EMIT Flu A samples. Dots of the same color indicate identical sequences, except for nodes colored gray.

## DISCUSSION

Our results, based on the available EMIT data, indicate that there is no strong evidence for compartment based differential population evolution in influenza A. We observe that the set of variants is mostly convergent within-participant over the course of infection post symptom onset. Variants differentiating between sample types are mostly specific to a single sample rather than common for all samples of a given type. Furthermore, we note that the SNPs that are sample type specific are mostly synonymous. Due to the lack of extensive epidemiological data, we were unable to investigate potential genomic markers for infection transmission routes, while the phylogenetic analysis suggests that samples analyzed in this study could be derived from a single introduction event. Our work suggests that further investigations into mutational spectra of influenza A are required to improve our understanding of the viral evolutionary processes.

There are multiple factors that could affect the sensitivity and specificity of intrahost variant calling in this study. Sequencing errors, PCR errors, and technical sequences within the sequencing data may lead to an increase in false positives variant calls (McCrone and Lauring 2016). Such errors could be largely reduced but not eliminated by the quality control process on the raw reads (Bolger et al. 2014; Chen et al. 2018; Xue and Bloom 2019) which we accordingly employed in our study. We have trimmed our reads to remove commonly known technical sequences, and discarded samples with very low or sparse coverage. This step is important since low read coverage on some of the regions in the reference genome may cause both sensitivity and specificity to decrease due to misidentification of true and false positive variants.

Another challenge in the intrahost variant calling process is the choice of an optimal minimum allele frequency threshold to identify variants with confidence (Maljkovic Berry et al. 2020). It is a trade-off between sensitivity and specificity where a higher threshold leads to an increase in specificity and a decrease in sensitivity, and vice versa. There is no single optimal minimum allele frequency threshold for calling intrahost variants, and the values used vary between the studies (Sobel Leonard et al. 2016; McCrone and Lauring 2016; Grubaugh et al. 2019) going as low as 1% and as high as 5%. In this study we used 2% as our cutoff which is within the range of commonly used values, as it allows us to stay sensitive to low frequency intrahost variants, while remaining reasonably above the error rates of the Illumina sequencing technology.

One of the main limiting factors of our current study is the lack of extensive longitudinal data for multiple study participants. In order to study more extensive viral intrahost evolution samples from prolonged infections (e.g. in immunocompromised hosts) will be required. Furthermore, investigating evolutionary processes and resulting mutational signatures of source environment requires experimental design that controls for potential confounders (Gutiérrez et al. 2012; Simmonds et al. 2019).

Another limiting factor in our study was lack of thorough epidemiological inter-personal interaction and exposure data that can aid in reconstruction of transmission clusters. Without epidemiological data, reconstruction of clusters and transmission pathways based purely on genomic data remains challenging (Frost et al. 2015; Wohl et al. 2016; Hill et al. 2021). In part, this can be explained by the narrow transmission bottleneck of the influenza A virus (Ghafari et al. 2020). This is further exacerbated by the varying viral load in different sample types, potential issues with amplification and sequencing, and previously outlined challenges in the intrahost variant calling. Although use of a Gesundheit-II bioaerosol sampler allows for direct collection of viral aerosols from human sources, the viral load detected in these samples is dependent on when samples were collected during the course of infection and individual characteristics of infected study participants (Yan et al. 2018). Compared with NPS data the geometric means and maxima for fine and coarse aerosols were 3-4 log_10_ RNA copies lower. Advancements in bioaerosol collection and recovery that could provide larger samples of aerosolized viruses could potentially lead to improvements in both RNA detection and depth of sequence coverage.

While our study on intrahost influenza evolution has unavoidable limitations, its explorative nature allows us to examine the mutational landscape of influenza A samples derived from different parts of participants’ respiratory tracts and the intrahost evolution of the virus in healthy hosts. This work on the available data shows that mutational signatures within samples are more specific to the participant than sample origin and are relatively stable for up to three days after the onset of symptoms. Although genomic data might aid the reconstruction of transmission events, distinguishing transmission pathways via genomic data alone remains challenging.

## Data Availability

Raw sequencing data and the variant calls used in this study are available at OSF (https://osf.io/bgt3n). Scripts used to process data and generate figures are available on GitLab: https://gitlab.com/treangenlab/reproduce/emit-scripts.

https://osf.io/bgt3n

https://gitlab.com/treangenlab/reproduce/emit-scripts.

## ACKNOWLEDGEMENTS

R.A.L.E. is supported by a training fellowship from the Gulf Coast Consortia, on the NLM Training Program in Biomedical Informatics & Data Science (T15LM007093). N.S., Y.L., and T.J.T were partially supported by funds from the Centers for Disease Control (CDC) contract 75D30121C11180 and P01-AI152999 NIH award.

EMIT was funded by the US CDC (Cooperative Agreement No. 1U01P000497-01). This work was also supported by the National Institute of Allergy and Infectious Diseases Centers of Excellence for Influenza Research and Surveillance (CEIRS) Grant Number HHSN272201400008C, and this funder had no role in study design, data collection and analysis, decision to publish, or preparation of the manuscript.

EMIT Investigators: Walt Adamson, Blanca Beato-Arribas, Werner Bischoff, William Booth, Simon Cauchemez, Sheryl Ehrman, Joanne Enstone, Neil Ferguson, John Forni, Anthony Gilbert, Michael Grantham, Lisa Grohskopf, Andrew Hayward, Michael Hewitt, Ashley Kang, Ben Killingley, Robert Lambkin-Williams, Alex Mann, Donald Milton, Jonathan Nguyen-Van-Tam, Catherine Noakes, John Oxford, Massimo Palmarini, Jovan Pantelic, Jennifer Wang (Scientific Advisory Board: Allan Bennett, Ben Cowling, Arnold Monto, Raymond Tellier)

